# Apple Watch and Withings Evaluation of Symptoms, Treatment, and Rhythm in those Undergoing Cardioversion (AWE STRUCk): A Pragmatic Randomized Controlled Trial

**DOI:** 10.1101/2021.07.10.21260230

**Authors:** Sanket S. Dhruva, Nilay D. Shah, Sreekanth Vemulapalli, Abhishek Deshmukh, Alexis L. Beatty, Ginger M. Gamble, James V. Freeman, James P. Hummel, Jonathan P. Piccini, Joseph G. Akar, Keondae R. Ervin, Kristine L. Arges, Lindsay Emanuel, Peter A. Noseworthy, Tiffany Hu, Victoria Bartlett, Joseph S. Ross

**Affiliations:** Department of Medicine, University of California, San Francisco School of Medicine, Section of Cardiology; Department of Medicine, San Francisco Veterans Affairs Health Care System, 4150 Clement St, Building 203, 111C, San Francisco, CA 94121; Division of Health Care Policy and Research, Department of Health Sciences Research, Mayo Clinic, Rochester, MN, USA; Robert D. and Patricia E. Kern Center for the Science of Health Care Delivery, Mayo Clinic, Rochester, MN; Duke Clinical Research Institute, Durham, NC, USA; Division of Cardiology, Duke University Medical Center, Durham, NC, USA; Department of Cardiovascular Medicine, Mayo Clinic, Rochester, MN, USA; Department of Epidemiology and Biostatistics, University of California, San Francisco School of Medicine; Department of Medicine, University of California, San Francisco School of Medicine; Center for Outcomes Research and Evaluation, Yale New Haven Hospital, New Haven, CT, USA; Section of Cardiovascular Medicine, Department of Internal Medicine, Yale University School of Medicine, New Haven, CT, USA; National Evaluation System for health Technology Coordinating Center (NESTcc), Medical Device, Innovation Consortium, Arlington, VA, USA; Yale University School of Medicine, New Haven, CT, USA; Section of General Internal Medicine, Yale School of Medicine, New Haven, CT, USA; Department of Health Policy and Management, Yale School of Public Health, New Haven, CT, USA

**Keywords:** medical devices, digital health, patient-reported outcomes

## Abstract

**Introduction:** Personal digital devices that provide health information, such as the Apple Watch, have developed an increasing array of cardiopulmonary tracking features which have received regulatory clearance and are directly marketed to consumers. Despite their widespread and increasing use, data about the impact of personal digital device use on patient-reported outcomes and healthcare utilization are sparse. Among a population of patients with atrial fibrillation undergoing cardioversion, our primary aim is to determine the impact of the heart rate measurement, irregular rhythm notification, and electrocardiogram features of the Apple Watch on quality of life and healthcare utilization.

**Methods and analysis:** We are conducting a prospective, open-label multicenter pragmatic randomized clinical trial, leveraging a unique patient-centered health data sharing platform for enrollment and follow-up. A total of 150 patients undergoing cardioversion for atrial fibrillation (or atrial flutter, if they have a history of atrial fibrillation) will be randomized 1:1 to receive the Apple Watch Series 6 or Withings Move at the time of cardioversion. The primary outcome is the difference in the Atrial Fibrillation Effect on QualiTy-of-life (AFEQT) global score at six months post-cardioversion. Secondary outcomes include inpatient and outpatient healthcare utilization. Additional secondary outcomes include a comparison of the Apple Watch ECG and pulse oximeter features with gold standard data obtained in routine clinical care settings.

**Ethics and dissemination:** The Institutional Review Boards at Yale University, Mayo Clinic, and Duke University Health System have approved the trial protocol. This trial will provide important data to policymakers, clinicians, and patients about the impact of the heart rate, irregular rhythm notification, and electrocardiogram features of widely used personal digital devices on patient quality of life and healthcare utilization. Findings will be disseminated to study participants, at professional society meetings, and in peer-reviewed journals.

**Trial registration number:** NCT04468321

**Strengths and limitations of this study:**

- This randomized clinical trial will use a patient-centered health data sharing platform for study enrollment and follow-up, leveraging patient ownership over health data and patient engagement with researchers.
- This is the first study to examine the impact of electrocardiogram and irregular rhythm notification features of personal digital technologies on patient-reported quality-of-life among patients with atrial fibrillation.
- Due to costs of personal digital devices, our study is powered to identify a difference of 8.8 on the primary outcome of Atrial Fibrillation Effect on QualiTy-of-life (AFEQT) global score, which is greater than the minimal clinically important difference of 5.
- Our study focuses on a subset of patients with atrial fibrillation, and the findings may not be generalizable to those who do not have atrial fibrillation.

## Introduction

The pace of innovation for digital health technologies is accelerating. Many individuals now have access to personal digital devices that track activity, sleep, and weight. Some devices also monitor physiologic measures such as oxygenation, heart rate and rhythm, and blood pressure.^1-3^ In response, the U.S. Food and Drug Administration (FDA) is working to establish a least burdensome regulatory framework for such technologies under its Digital Health Innovation Action Plan.^4 5^ For software-based medical devices, the agency has initiated a Software Precertification (Pre-Cert) Pilot; a key element is collecting and interpreting post-market, real-world data about software performance.^6^ In September 2018, FDA cleared 2 software features for the Apple Watch (Apple, Cupertino, CA) through the De Novo medical device regulatory pathway: 1) to detect irregular heart rhythms likely to be atrial fibrillation^7^ and 2) to generate a single-lead ECG.^8^ However, several risks were identified, including: misinterpretation and/or over-reliance on the device, false negative results, and false positive results. Strategies to mitigate these risks were also identified.^7 8^ Additionally, multiple other digital devices have come to market that can perform ECGs and offer irregular rhythm notifications. Since that time, Apple has released an oxygen saturation monitor, which was not cleared by the FDA.

Catalyzed by the coronavirus disease 2019 (COVID-19) pandemic, these digital health monitoring technologies have been rapidly integrated into clinical practice to support patient monitoring in lieu of in-person visits, thereby reducing possible patient and clinician exposure.^9^ Insurers also sometimes provide coverage for these devices. ^10^ These devices have inverted the traditional pyramid of scientific discovery.^11^ Medical products (i.e., traditional new medical devices and drugs) are generally discovered and tested through scientific research that leads to regulatory approval and is disseminated to clinicians, who in turn inform patients of tests and treatments authorized for commercial use. While traditional medical devices and drugs can be marketed directly to patients, clinician prescriptions or clinical care are necessary for patient use.

In contrast, personal digital devices follow a different path. Many (or all) health and wellness-focused features are not reviewed or regulated by the FDA, while others are cleared if they are similar to previously cleared or approved medical devices. They are directly marketed to the public by manufacturers; patients consume marketing information and may purchase and use the devices without clinician oversight – or any direct interaction with the medical community. Patients may approach clinicians about using personal digital devices, but clinicians may not have scientific research available to guide them.^11^ This issue occurs with the Apple Watch, which is widely advertised and now exceeds 30 million units in annual sales. However, there are limited data on the impact of commonly used health features such as heart rate measurement, irregular rhythm notification and electrocardiogram (ECG) features on patient quality of life^12^ and healthcare utilization. Further, there have been limited evaluations of the accuracy of more novel health features such as the single lead ECG and pulse oximeter features.

Therefore, there is a need to assess the clinical impact of these devices through active surveillance to guide future labeling and risk mitigation strategies. While the Apple Heart Study reported positive predictive values for the irregular pulse notification^2^ and a subsequent study of cardiac surgery patients provided data on accuracy of atrial fibrillation detection,^13^ it is unknown how such notifications affect patients. There has been concern expressed about smartwatches leading to patient anxiety.^14^ In fact, anxiety disorders associated with these devices have been documented.^15^ Further, there are limited data about the impact on healthcare utilization of Apple Watch heart rate, irregular rhythm notification, and ECG features. One single-health system study found higher overall healthcare utilization among patients with atrial fibrillation using personal digital devices, including the Apple Watch, as documented in clinical notes compared to a propensity-matched sample of patients without documentation of these devices; the authors concluded by calling for prospective, longitudinal, randomized data on health outcomes and healthcare use.^16^ Some clinicians have expressed worry about cascades of testing to confirm or rule out disease.^14^ One single-center study found that only 11.4% of patients had a clinically actionable diagnosis of interest after an irregular rhythm notification, although several tests (including Holter monitors, echocardiograms, and even computed tomography scans of the chest) were performed.^17^ Additionally, a small study has raised the possibility that Apple Watch heart rate measurements among patients with atrial fibrillation may sometimes be inaccurate;^18^ this is a known limitation of photoplethysmography-based heart rate monitors, given pulse deficit can occur in atrial fibrillation.^1^ Therefore, further validation is needed.^18^

Finally, there is a need to evaluate the accuracy of the newer ECG and pulse oximeter features. The ECG feature has been compared to 12-lead ECGs in 2 studies, although one focused only on the QT interval in sinus rhythm^19^ and another included only healthy volunteers.^20^ There are no published data about the accuracy of the oxygen saturation monitor.

Accordingly, we designed the **A**pple Watch and **W**ithings **E**valuation of **S**ymptoms, **T**reatment, and **R**hythm in those **U**ndergoing **C**ardioversion (AWE STRUCk) trial, a pragmatic randomized controlled trial (RCT), to address these issues. This trial will provide an independent assessment of the impact of the Apple Watch heart rate, irregular rhythm notification and ECG features on 1) quality of life, 2) healthcare utilization, and 3) accuracy of measurements among patients undergoing cardioversion for atrial fibrillation at three geographically distinct academic medical centers in the U.S. By comparing the Apple Watch to a personal digital device without the ability to perform an ECG, offer irregular rhythm notifications, or measure pulse oximetry, we week to better understand the impact of these health features of the Apple Watch. In addition to informing physicians, patients, and policymakers, these data can inform future labeling and risk mitigation strategies that could impact millions of individuals.

### Overall Study Design

AWE STRUCk is a prospective, open-label multi-center pragmatic RCT designed to evaluate the impact of patient use of the Apple Watch, focused on its accompanying health features of heart rate measurement, irregular rhythm notification, and electrocardiogram features on patient-reported outcomes and healthcare utilization at six months. Patients with a history of atrial fibrillation who are scheduled for cardioversion are randomized 1:1 to receive the Apple Watch Series 6 or the Withings Move (**Figure 1**). Patients and the public were not involved in study design.

**Figure 1:**
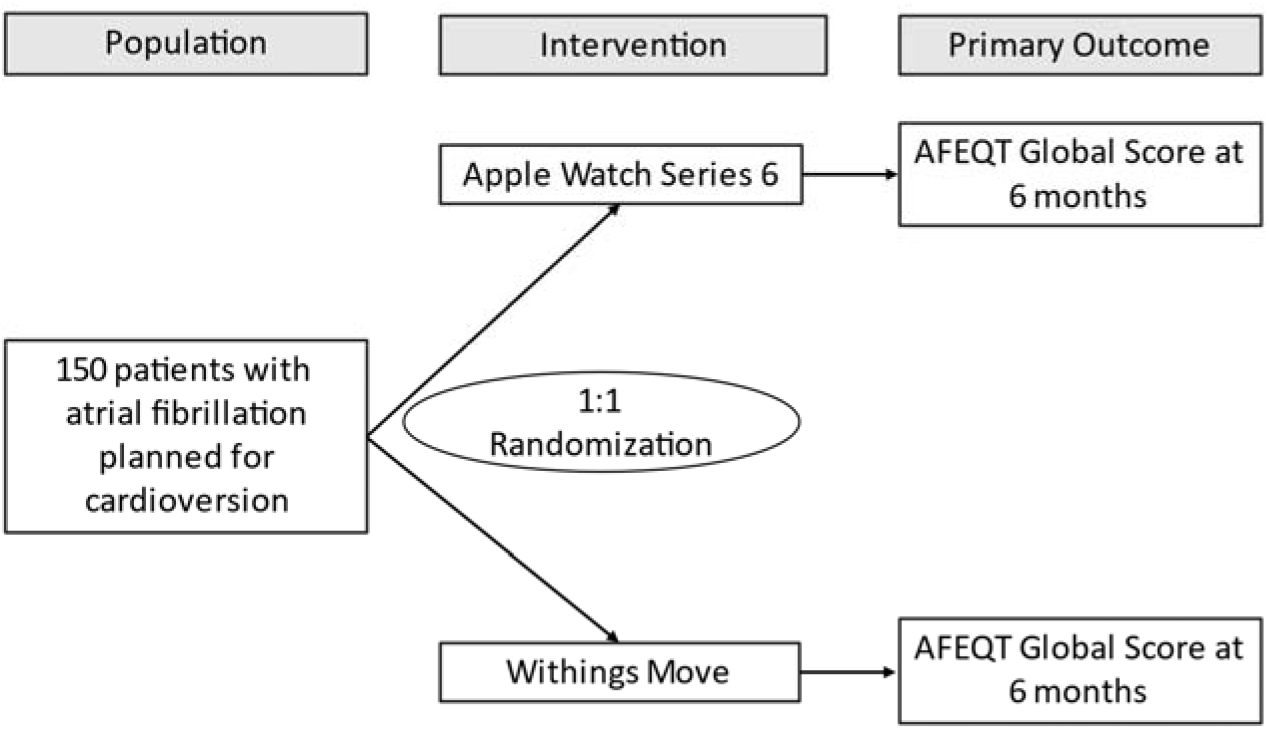
Study Flow Diagram

### Data Acquisition

AWE STRUCk leverages a unique patient-centered health data sharing platform, Hugo, which is intended to overcome many of the limitations of traditional clinical trials.^21^ Hugo aggregates electronic health data for each patient from multiple sources, including electronic health records from hospitals and physicians’ offices, along with data from personal digital devices, by leveraging Blue Button technology and Application Programming Interfaces (**Figure 2**). Blue Button technology refers to functionality that enables individuals to download their health records.^22^ Hugo also supports direct patient surveys through text messaging or emails, enabling assessment of patient-reported outcomes and other information without requiring patients to meet directly with study coordinators after initial enrollment. All participants will be enrolled in this platform, through which they will receive near real-time access to their electronic health data from these multiple sources and then share these data with researchers.^23^ No data, therefore, are directly obtained from health systems.

**Figure 2:**
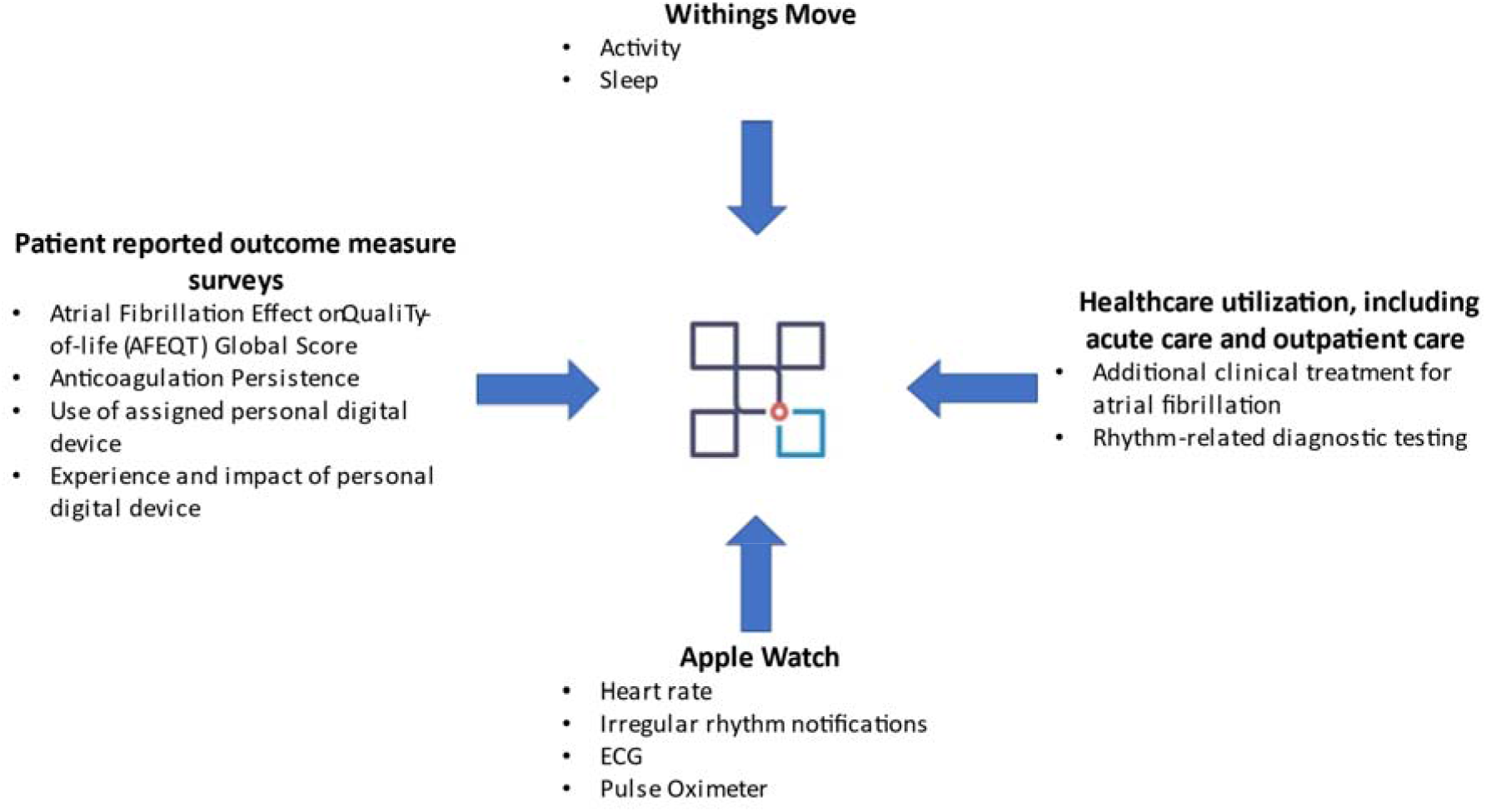
Data Aggregated Using the Hugo Platform

### Sample Selection

A total of 150 patients will be enrolled at one of three academic health systems in equal numbers (50 at each health system): Yale New Haven Health, Mayo Clinic, and Duke Health. Potential patients will be identified by members of the clinical care team if they have been referred for cardioversion for atrial fibrillation (or atrial flutter, if they have a history of atrial fibrillation). If a patient in the outpatient setting expresses interest in the study and meets inclusion criteria of having an iPhone and receiving both primary and cardiology care at one of the academic health systems, then a research team member will follow-up with them to verify additional inclusion criteria and ask the patient to arrive early for enrollment prior to their cardioversion. Patient identification will vary slightly by site, but essentially hospitalized patients planned for cardioversion will be identified prior to their procedure and, if they meet inclusion criteria, will be offered the opportunity to enroll.

### Inclusion and Exclusion Criteria

Patients must fulfill the following criteria prior to trial enrollment:

1. Age >22 years at the time of enrollment
2. English-speaking
3. Planned for direct current cardioversion for atrial fibrillation (or atrial flutter, if they have a history of atrial fibrillation), as noted by referring clinical staff or on chart review
4. Lives independently and does not require continuous care
5. Has an email account (or is willing to create one)
6. Has a compatible smartphone (iPhone 6s or later)
7. Willing to wear only the device to which they are randomized to receive for the study period for as many hours during the day as able, except for time spent charging the device or in environments that may be suboptimal for the device
8. Willing to use the Hugo Health platform
9. Receives both primary and cardiology care at one of the study centers

#### Exclusion criteria

Patients with any of the following will not be eligible for trial enrollment:

1. No history of atrial fibrillation
2. Not able to independently adhere to study protocol, including read and sign consent
3. Enrollment in another study protocol

Inclusion criteria will be confirmed prior to the informed consent process and documented. Patients who do not meet all inclusion criteria or who meet any of the exclusion criteria will not proceed with consent and enrollment. Study coordinators will obtain informed consent, and patients will sign a digital consent form (**Supplemental Material**).

### Interventions

After signing informed consent documentation, patients will be randomized 1:1 to the Apple Watch Series 6 (GPS) or Withings Move arm using a central computer-generated randomization algorithm that is stratified by site to ensure balance. We will pre-specify that each of the three clinical sites will enroll a total of approximately 25 patients in each arm.

The Apple Watch Series 6 is a smartwatch manufactured by Apple (Cupertino, CA) that can obtain multiple physiologic measurements, including activity, sleep, and several cardiopulmonary parameters. Activity features include steps taken, flights of stairs climbed, walking and running distance, energy expenditure, and data about time spent standing. Cardiac features include heart rate, both at rest and with activity, and heart rate variability. Additionally, the Apple Watch offers optional notifications for low and high heart rates and irregular rhythms. The triggers for low and high heart rate notifications can be customized by the user. The device also allows users to perform a single-lead ECG and pulse oximetry measurement. The Withings Move is a smartwatch manufactured by Withings (France) that enables tracking of activity and sleep. Activity features include steps taken, distance traveled, and energy expenditure. Sleep features include details about light and deep sleep phases, as well as sleep interruptions.

For this study, all data will be confidentially aggregated using Hugo. Patients randomized to the Apple Watch arm will link their Apple HealthKit account to the Hugo platform, which will enable receipt of Apple Watch-related data. Patients randomized to the Withings Move will share data using a Withings Health Mate account, which will also be linked to the Hugo platform. Research coordinators at each clinical site will assist patients with creating and linking these accounts. Patients who would like to do so will also be able to share their pharmacy data and Medicare claims data. Patient-reported outcome measure (PROM) questionnaires will be texted or emailed to patients (depending on patient choice) and will enable patients to answer queries using a secure link. Patients will be reminded to wear their personal digital device and share data, as well as to complete PROM questionnaires in order to improve adherence and response rates. Patients will be able to discontinue study participation at any time during the study.

### Compensation

All patients will be able to keep the personal digital device to which they have been randomized: the Apple Watch Series 6 (GPS), with fair market value $399 in mid-2021 and the Withings Move with fair market value $70. Additionally, patients randomized to the Withings Move will receive a stipend for time contributed to study participation and survey completion because of the lower cost of this device. The estimated hourly stipend is based on the average minimum wage across the recruitment sites (approximately $10), resulting in a total $80 stipend. Patients will receive payments every 30 days upon completing surveys; these payments will be provided as a digital Visa card.

### Demographic and Clinical Characteristics (Table 1)

Demographic characteristics to be collected at enrollment include age, sex, race, and ethnicity through patient self-report with discrete response categories. Patients will also self-report the presence of multiple cardiovascular and non-cardiovascular comorbidities through a digital questionnaire at enrollment.

**Table 1.** Demographic and Clinical Variables Obtained at Baseline from Patient Self-Report.

| **Demographics** | **Clinical Comorbidities** |
| --- | --- |
| Age | Hypertension, also called high blood pressure |
| Sex | High cholesterol |
| Race | Coronary heart disease |
| Ethnicity | A heart attack (also called myocardial infarction) |
|  | A stroke or transient ischemic attack (TIA) |
|  | Emphysema |
|  | Asthma |
|  | Cancer or a malignancy of any kind |
|  | Diabetes |
|  | Weak or failing kidneys |
|  | Any liver condition |
|  | Post-Traumatic Stress Disorder, or PTSD |
|  | Depression |
|  | General Anxiety Disorder |

### Outcomes (Table 2)

#### Primary Outcome

Nearly all data elements for outcomes will be aggregated through the Hugo platform. The primary outcome is the difference in the Atrial Fibrillation Effect on QualiTy-of-life (AFEQT) questionnaire^24^ global score at six months compared to baseline. The AFEQT questionnaire is a validated, 20-item questionnaire that has been shown to reliably track clinical changes in patients with atrial fibrillation. It contains two sections. The first section asks about the last episode of atrial fibrillation (one question). The second section contains 19 questions in four domains: symptoms (four items), daily activities (eight items), treatment concern (six items), and treatment satisfaction (one item). A composite overall score that ranges from 0 to 100 is calculated from the symptoms, daily activities, and treatment concern domains; higher values suggest better quality of life. Each domain also has a corresponding subscale score that also ranges from 0 to 100. Published data indicate that a change of approximately five in the overall score indicates a clinically meaningful difference.^25^ We will modify the AFEQT questionnaire into a digital format.

#### Secondary Outcomes

The secondary outcomes can be divided into three distinct groups (**Table 2**). The first group of secondary outcomes includes five patient-reported outcomes. First, we will examine the change in AFEQT global score at 1, 2, 3, 4, 5, and 12 months (the primary outcome examines this score at 6 months); this will provide additional insight into changes in the primary outcome. Second, at all time points we will examine the change within individual domains of the AFEQT questionnaire (symptoms, daily activities, treatment concern, and treatment satisfaction); this will allow us to determine if the personal digital devices have different effects on specific patient-reported outcomes. Third, we will ask about anticoagulation persistence at 1, 2, 3, 4, 5, and 6 months and again at 12 months, since clinical practice guidelines recommend that patients receiving cardioversion should be on therapeutic anticoagulation for at least 4 weeks post-cardioversion,^26^ and often patients meet criteria for longer or indefinite anticoagulation; we will determine this through adopting the Brief Medication Questionnaire and asking patients if they have taken their anticoagulant, and for how many days in the past 1 week. Fourth, we will ask patients at 1, 2, 3, 4, 5, and 6 months and again at 12 months if they have stopped using the personal digital device to which they were assigned (and, if so, the reason); if they have used a different personal digital device (and which device). Fifth, we will ask patients at 1, 2, 3, 4, 5, and 6 months and again at 12 months if they received any abnormal or worrisome notifications from their wearable device and, if so, if they contacted their physician and the result of the contact. Among patients randomized to the Apple Watch, we will also ask if they have used the device to examine heart rhythm for symptoms. Sixth, we will ask at 6 and 12 months about patient experience with technology; reassurance or worry about overall health; reassurance or worry about atrial fibrillation; and confidence about atrial fibrillation, physical activity, and usual activities. Patients randomized to the Apple Watch arm will also be asked about the amount of notifications.

The second group of secondary outcomes are healthcare utilization outcomes. There are four healthcare utilization outcomes, all of which seek to determine if patients randomized to the Apple Watch (with cardiopulmonary features) have different rates of obtaining care, including diagnostic testing and clinical treatment for atrial tachyarrhythmias compared to patients receiving the Withings Move. First, we will determine if patients receive additional clinical treatment for atrial fibrillation or flutter at 6 and 12 months by examining a composite of rhythm control intervention, which is defined as at least one additional cardioversion, initiation of antiarrhythmic medical therapy, dose escalation of antiarrhythmic medication, change to another antiarrhythmic medication, or ablation for atrial fibrillation. The medication data will be obtained primarily from electronic health record data. Second, we will compare acute care use, a composite of emergency department visits, observation stays, and all hospitalizations. Third, we will compare outpatient care use, a composite of outpatient primary care visits, outpatient cardiology or cardiac electrophysiology visits, and scheduled telephone encounters. Fourth, we will compare rhythm-related diagnostic testing at 6 and 12 months, a composite of total ECGs and total outpatient heart rhythm monitors.

**Table 2.**
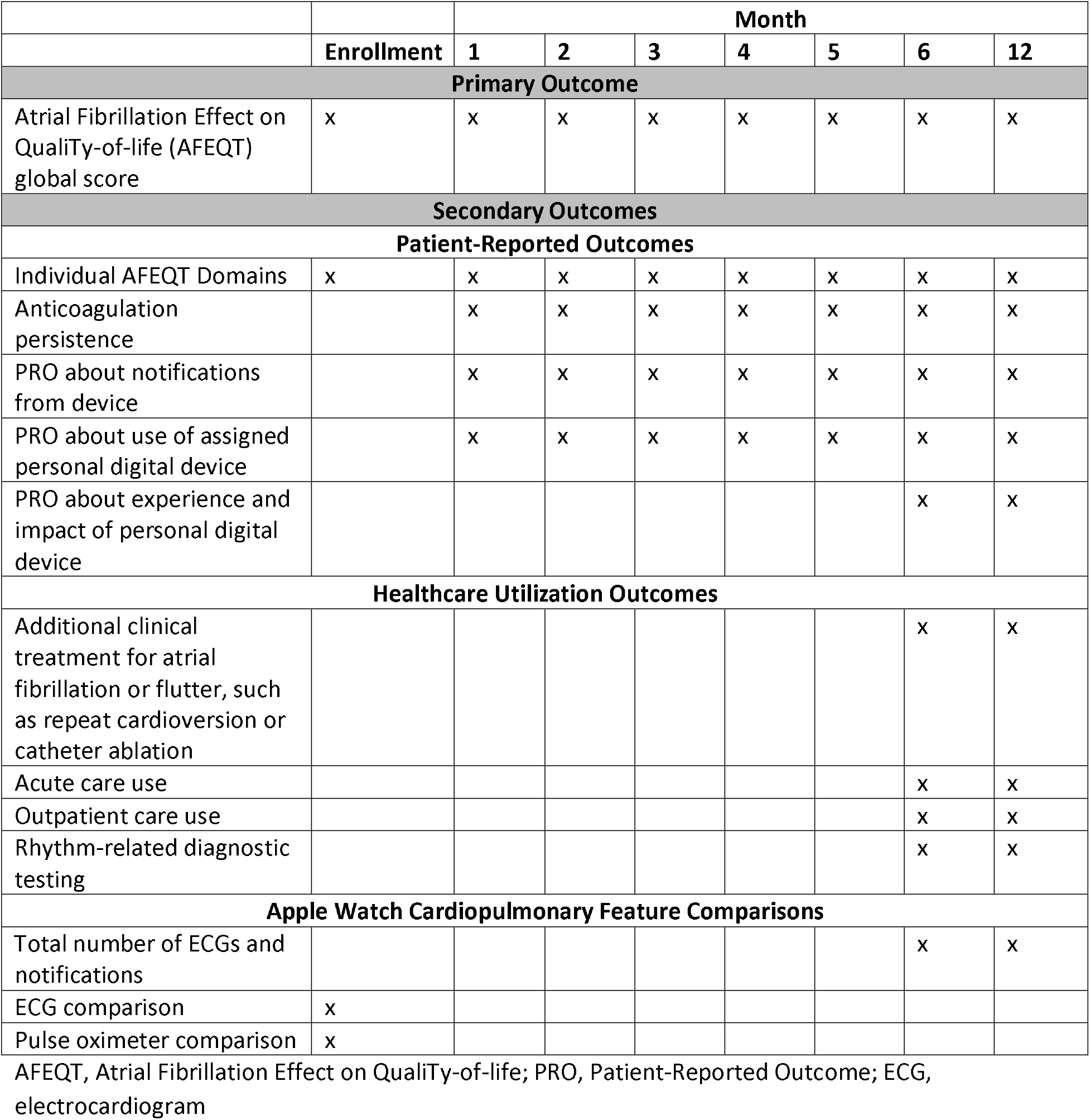
Primary and Secondary Outcomes.

The third group of secondary outcomes are only determined among patients randomized to the Apple Watch. We will determine the total number of notifications for irregular rhythm, high, and low heart rates. To address the knowledge gap about the accuracy of the Apple Watch ECG, we will perform a comparison between each Apple Watch ECG (using 10-second interval with highest resolution and free from artifact) vs 12-lead ECG obtained in routine clinical practice as close to simultaneous as possible, ideally within a maximum window of 30 minutes to the Apple Watch ECG. The parameters that will be compared include heart rhythm (identical to 12-lead ECG or different), rate (difference in beats per minute), and intervals: PR, QRS, and QT (difference in milliseconds). These metrics will include both inter-observer and intra-observer reliability. The Apple Watch ECG will be obtained by asking each patient to email their Apple Watch ECG(s) to a secure study team email address at each health system, while information on the number of ECGs will be aggregated through Hugo. Finally, we will compare pulse oximeter readings from the Apple Watch Blood Oxygen measurement to a standard, medical grade pulse oximeter in clinical practice. The pulse oximeter data will be obtained when a research coordinator is at the patient’s bedside during or after enrollment and attaches a medical grade pulse oximeter; the values will be entered by a patient into a survey when the research coordinator is present.

#### Sample Size Calculation

Our sample size was determined assuming 80% power to detect a differential change of 8.8 on the AFEQT questionnaire global score at 6 months, the primary outcome, with alpha 0.05. This effect size is greater than the expected minimal clinically important difference of 5.^25^ This estimate accounts for an estimated drop-out rate of 8% with respect to non-completion of the AFEQT questionnaire at 6 months.

For the secondary endpoints focused on healthcare utilization, we expect drop-out to be close to zero because patients are followed passively with minimal burden and receive both primary and cardiology care at the enrolling health system. Additionally, the Hugo platform enables patients to share data from other health systems. Therefore, even if a patient switches primary or cardiology care to outside one of the health systems of interest, if he/she connects their new health system (as portal availability is legally required), comprehensive outcome ascertainment will remain possible.

Validation of Apple Watch ECG readings with a simultaneous 12-lead ECG in clinical practice will be performed for as many ECGs as qualify. Similarly, pulse oximeter readings will be performed and validated against those measured using a clinical pulse oximeter whenever possible.

#### Data Analysis Plan

Data quality and integrity checks will be performed. Baseline descriptive statistics will be reported for the overall sample, and for both the Withings and Apple Watch arms. These will include patient age, sex, race, ethnicity, and the comorbidities collected. Baseline data will be compared by Chi Square (or Fischer’s Exact) test for dichotomous/categorical variables and t-tests for continuous variables and the median test for non-parametric variables. Data on missing covariates will be missing and not imputed.

For the primary outcome, we will calculate the difference in the AFEQT global questionnaire score (which ranges from 0 to 100) at baseline and at 6 months, and compare between patients randomized to the two study arms using a t-test.

The secondary outcomes will use a t-test for comparison of the AFEQT global questionnaire score between baseline and at 12 months and a t-test also for the individual domains at 1, 2, 3, 4, 5, 6, and 12 months. The secondary healthcare utilization outcomes are composites and will use a t-test or the Wilcoxon rank-sum test for comparison between the two study arms. For the ECG comparison, inter-observer and intra-observer agreement will be assessed using intraclass correlation coefficients. In an exploratory analysis, the difference between the readings on the clinical pulse oximeter and Apple Watch pulse oximetry feature will be compared.

If patient drop-out or loss to follow-up occurs, then we will carry forward the last patient-reported outcome measure response. If an electronic health record data connection is lost for a study participant, we will only include the follow-up duration when data were available in the utilization endpoint analyses. All p-values will be significant at <0.05 with two-sided inferential tests. All analyses will be conducted at Mayo Clinic.

### Ethics and Dissemination

#### Ethics Approval

The AWE STRUCk RCT is sponsored by the National Evaluation System for health Technology Coordinating Center (NESTcc). Ethics approval was obtained independently at each of the 3 health systems, including at Yale University on August 2, 2020; at Mayo Clinic on March 3, 2021; and at Duke University Health System on June 4, 2021. Any amendments to the protocol are first reviewed by each of the three local institutional review boards prior to implementation and also receive approval from the study sponsor. This RCT is also registered at ClinicalTrials.gov (NCT04468321) and was first posted on July 13, 2020.

Serious adverse events are not expected in this study of consumer personal digital devices. However, if there are device-related adverse events, they will be reported to the FDA’s Manufacturer and User Facility Device Experience (MAUDE) database. A data safety monitoring board was not convened because the intervention is low-risk.

The investigator team will make clear that any sync-able data, including PROMs, will not be reviewed in real-time by researchers and will not be provided to the clinical care team and, therefore, any adverse or severe symptoms should be reported directly to their physician(s) or emergency room physicians as they would have in the normal course of care.

#### Dissemination

RCT results will be presented at both scientific meetings and submitted for publication to peer-reviewed journals. Additionally, study results will be shared with enrolled study participants.

## Discussion

The AWE STRUCk pragmatic RCT will address important knowledge gaps about the impact of novel, widely used personal digital devices on quality of life and healthcare utilization. Despite the widespread and increasing use of these devices for both consumer and clinical applications, few studies have assessed their impact on these important outcomes. Some of the features are also marketed as health and wellness, without a requirement for FDA clearance. To our knowledge, no prospective studies of these technologies focused on these endpoints have been pursued independent of the device manufacturers; additionally, manufacturers have not conducted many studies in these areas and may lack incentives to do so.

In addition to the data generated, there are several strengths to this study. First, the use of a controlled design helps to isolate the impact of the cardiopulmonary features (heart rate, ECG, irregular rhythm notification, and pulse oximeter) from activity tracking. Second, this study employs a novel pragmatic digital paradigm of research in which patients partner with researchers to share their data; data do not come directly from traditional sources, such as health systems. This research paradigm is supported by the principle of individual agency over data.^21^ Third, recent policy changes by Medicare provide reimbursement for remote patient monitoring using personal digital devices and this study will provide information about the implications of these changes.^27 28^

This RCT should also be considered in the context of multiple possible limitations. First, the study is powered to identify a difference of 8.8 in the AFEQT global score, which is greater than the minimal clinically important difference of 5,^25^ due to the high costs of the devices provided to patients and the budgetary constraints of the trial. Second, the PROMs have been digitally adapted. The AFEQT questionnaire has been validated in paper format.^24^ However, the vast majority of PROMs that are adapted to a digital format result in synchronous responses^29^ and this approach is more accurate than clinician documentation.^30^ Third, there is the possibility of incomplete ascertainment of events if patients use a different health system for care and do not connect these with Hugo. However, the Hugo platform allows patients to add data from multiple health systems (and other sources) at any time throughout the study, thereby facilitating complete clinical event ascertainment. Fourth, the irregular rhythm notification is not intended for use in individuals previously diagnosed with atrial fibrillation. However, the ECG feature is not recommended for users with arrhythmias *except* for atrial fibrillation and sinus rhythm (since the software can only distinguish these two heart rhythms)^8^. People with atrial fibrillation may be more likely to purchase this device than others to monitor their rhythm and correlate with symptoms. Indeed, in the Apple Heart Study, nearly 20% of first study visit participants reported a diagnosis of atrial fibrillation/flutter before enrollment, even though a history of atrial fibrillation was an exclusion criterion.^2^ Few digital health studies have enrolled patients with clinical morbidities to examine outcomes; most studies have instead focused on healthy volunteers.^31^ Our study will address this limitation. Additionally, a study examining the impact of the Apple Watch on patient-reported outcomes and healthcare utilization requires an enriched population; otherwise, study duration would be many years with a population size multiple times larger than our trial, resulting in significant cost and delays in generating much-needed evidence.

### Data Sharing Plan

The study protocol, statistical analysis plan, informed consent form, and analytic code will be publicly available. After study completion, our intention is to share de-identified individual patient data for use by others for research in a manner that protects patient confidentiality.

## Supporting information

SPRIT Checklist

Informed Consent Form: Yale University School of Medicine

## Acknowledgments

We would like to thank Harlan M. Krumholz, MD, SM, for his guidance on the conception and design of this trial and Jessica Ritchie and Laura Ciaccio for their assistance in the design and implementation of this trial.

## Contributors

SD, ND, SV, AD, AB, GG, JF, JH, JP, JA, PN, JR designed the study. SD and JR wrote the first draft of the manuscript. GG is the overall study coordinator and TH, KA, and LE are individual site study coordinators. All authors revised the manuscript and approved the final version of the submitted manuscript. The corresponding author attests that all listed authors meet authorship criteria and that no others meeting the criteria have been omitted. SD acts as the guarantor.

