## Supplementary material for "Apple Watch and Withings Evaluation of Symptoms, Treatment, and Rhythm in those Undergoing Cardioversion (AWE STRUCk): A Pragmatic Randomized Controlled Trial": Informed Consent Form: Yale University School of Medicine

### COMPOUND AUTHORIZATION AND CONSENT FOR PARTICIPATION IN A RESEARCH PROJECT
# 200 FR. 4 (2016-2)
### YALE UNIVERSITY SCHOOL OF MEDICINE – YALE-NEW HAVEN HOSPITAL

**Study Title:** Effect of Wearable Devices on Patient-Reported Outcomes and Clinical Utilization: A Randomized, Controlled Trial

**Principal Investigator:** **Joseph Ross, MD, MHS**
Professor of Medicine (General Medicine)
Yale School of Medicine
Best Contact Number: 203-785-2987

**Funding Source:** National Evaluation System for health Technology Coordinating Center (NESTcc)

#### Key Summary of Research

You are invited to take part in a 12-month research study that will follow patients undergoing cardioversion for atrial fibrillation at Yale-New Haven Hospital. The study uses a technology platform that, with your permission, will gather information from your online health records. This technology platform also collects your responses to questionnaires from the researchers conducting this study. Finally, it will collect information from a smartwatch that we will provide you to keep. You may need to download an app to link your device and view your health information that is aggregated by this platform. Your participation is entirely voluntary, and your clinical care will not be affected based on whether or not you choose to participate. If you choose to participate, you can change your mind at any time and withdraw from the study. You will not be penalized in any way or lose any benefits to which you would otherwise be entitled if you choose not to participate in this study or if you choose to withdraw.

For you to decide whether you want to be part of this study, we want to make sure you know enough about the study risks and benefits to make an informed decision. This consent form gives you detailed information about the study, which a member of the research team will discuss with you. This discussion will review all aspects of this research, especially the confidentiality risks of you having personal health information on your mobile device should you need to download the app. Once you understand the study, you will be asked if you want to participate; if so, you will sign this form electronically and be sent a digital copy of the document for your records.

This study uses a technology platform called Hugo that you can access on your mobile phone or other devices that are connected to the internet. Hugo collects data from multiple sources to create your own personal health record (PHR) and empowers you to share that data with researchers. You will have control over which data sources you connect to Hugo and will have the option to turn off data sharing at any time. Hugo has a one-way link to your health care providers: it can access information but cannot add any information to your health records.

The purpose of this study is to help researchers better understand the impact of wearable devices on patient experience and clinical trends following cardioversion. We want to know if wearing one of these devices changes the way you feel following your procedure or your accessing medical care.

You have been invited to join this study because you will be undergoing a cardioversion procedure for atrial fibrillation at Yale-New Haven Hospital. If you agree to take part in this study, you will be one of 50 patients participating in this study in the Yale-New Haven Health System. Patients must be 22 years of age or older and have a compatible smartphone with access to the internet. You will be asked to connect your electronic medical records from your Yale-New Haven Health MyChart account, electronic medical records from any other health systems where you receive care, information from iPhone health application(s), and information from a wearable device that we will provide to you, to a personal health record platform called Hugo. Through Hugo, you will also be asked to answer short questionnaires that will be sent to you via your choice of email or text monthly for 6 months after your procedure and then also at 12 months after your procedure to better understand your health outcomes.

With your permission, the researchers will be able to view your data soon after you visit a doctor without asking you to keep track of this information yourself. In addition, the data from your connected wearable device and from the questionnaires you complete will feed into the Hugo platform. This allows researchers to access your information in one location, so they can better understand your experience following your procedure. You will receive one of two possible wearable devices for this study, both will have the ability to monitor your activity levels and sleep, with one having additional cardiac features. We will provide you with the wearable device, and it will be yours to keep after the study is over.

All research studies involve some risk to participants. For this study, you are sharing your personal health information with researchers and there is a risk that someone might access that information improperly. The research team has strict protocols in place to control access to your data, including keeping audit logs of who has accessed your data via Hugo. Researchers will only be able to view the health data that you connect to the Hugo platform, and you will be able to withdraw from the study and stop sharing data through Hugo at any time. You can stop sharing your data by disconnecting your portals within the Hugo platform under the connected portals menu or by reaching out to a study coordinator. The researchers will not give anyone, including your doctors, any information from the questionnaires you fill out; that data will be kept confidential and will not be part of your medical record.

#### Description of Procedures

*Setup process*

If you agree to take part in this study, the initial set up process could take approximately 1 hour in total. You will be asked to take part in the following steps:

1. Using your own mobile device or a laptop available through the research team, a research associate working with the research team will help you register for the Hugo platform; you may need to download the app. Registration for Hugo will require you to enter basic information including first name, last name, email address, and to choose a password that will be known only to you. You will then be prompted to accept the standard terms and conditions as well as a privacy notice for the Hugo platform.

2. Using your personal mobile device (phone or tablet), you will check your email and click the confirmation link to activate your new Hugo account. If you are unable to access your email on your own mobile device, a laptop will be available for you to use. If you do not have an email account and wish to create one, the research associate can help you set one up from a variety of free email providers.

3. Once your Hugo account is confirmed, the research associate will then walk you through the remaining steps to complete study enrollment in the Hugo platform.

4. The research associate will show you how to access and complete your enrollment questionnaire. You can choose whether this questionnaire will be sent to you through email or text message. The email or text message will securely link to a multiple-choice survey in a web browser. The research associate will help you with any technical questions you may have when you begin the survey.

5. The research associate will then provide you with a wearable device to which you have been randomized, which you will use throughout the study. They will help you set up an account for this device, including connecting it to both your phone and the Hugo platform.

    a. Should you receive a device that allows you to track cardiac symptoms, you will be asked to take 2 types of readings before and after your procedure. The research associate will show you how to do so.

6. The research associate will assist you in linking your iPhone health application(s) to the Hugo platform.

***Please note:*** Researchers will **not be watching or evaluating your symptoms as part of this study,** including your responses to the questionnaires. None of the information collected in this study will be shared with your medical team. If at any point you begin to experience new symptoms, or any medical issues arise, **please contact your doctor. In case of a life-threatening emergency, call 911 immediately.**

**Should there not be enough time to complete all enrollment steps before your scheduled procedure time, the remaining steps may be completed post-procedure*

7. The Hugo platform will prompt you to link your patient portal accounts by presenting a list of participating health systems. You can select the systems where you have received care and enter your patient portal login and password. If you have forgotten your password, you can request a reset link be sent to your email account. The research associate can assist in setting up a new YNHH MyChart account, obtain your YNHH MyChart username, and help reset your YNHH MyChart password if needed.

8. The research coordinator can help you set up accounts for other health systems and pharmacies, including CVS, Walgreens, or Walmart.

9. After your health records have been linked, the Hugo platform will display your health data. The research associate will help you with the study information and be available to answer any questions related to data sharing.

10. If you have health insurance coverage through Medicare (available to those 65 years or older and younger people with disabilities and people with End-Stage Renal Disease), the research associate can also help you connect to your Centers for Medicare and Medicaid Services (CMS) Blue Button account. Blue Button gives you access to your Medicare claims data. With your permission, Hugo can gather information from your claims data to provide information on the health care you receive while participating in the study.

11. At the end of this consent form, we will ask you to give the researchers permission to see the health information and survey responses that you connect or provide to the Hugo platform. The medical record data being shared will be what is available electronically via the patient portal and may include Medications, Problems, Allergies, Procedures, Encounters, Lab Results, Diagnoses, Vital Signs, Immunizations, and possibly other data that becomes available. From iPhone health application(s) and your wearable device, the data being shared may include sleep patterns, movement (steps per-day), heart rate, rhythm notifications, pulse oximetry, weight, BMI, and other available data.

*Continuous Study Process*

After the initial in-office set up is complete, you will be asked to perform the following tasks at home. If you have any questions or experience technical issues at any time, please reach out to the study team via email at:

- For the study, we ask that you wear your provided device as often as possible both during the day and while sleeping. We also ask that you leave your Bluetooth turned on during the study period. If you are unfamiliar with the Bluetooth setting on your phone, the research associate will be able to help you. We also ask that you do not share your iPhone or iCloud account with anyone else during this time.

    - If you do not use your provided device for 7 consecutive days, you will be sent a reminder (text or email, based on your preference). If no data are received within 24 hours, then you will receive up to two additional reminders. If no data are received within an additional 48 hours, a research associate will reach out to you via phone to follow-up.

- Should you receive a device that allows you to obtain an ECG, we ask that you use your device to record an ECG/EKG anytime, at approximately the same time, you have one recorded as part of your normal care. We also request that you then email this ECG to us. This will enable us to compare the ECG to the one that you receive during normal care.

- Surveys will be sent to you every 30 days throughout the duration of the follow-up period in addition to a survey at 12-months post-procedure
    - You will be sent reminders to complete these surveys at 24 hours if no response is received and again at 48 hours if there is still no response
    - If after an additional 48 hours no survey response is captured, a research associate will attempt to contact you via phone to determine if there are any technical issues.

- We estimate that your participation in the study, including completing the enrollment and set-up process and answering the questionnaires should take you no more than 8 hours over the study.

**New Information**

You will be informed of anything that happens during the study that may cause you to re-think your decision to continue participation.

**Risks and Inconveniences**

The primary risk is loss of confidentiality and privacy. The risk to your privacy is that the Hugo platform collects personally identifiable information (like your name and where you go to the doctor) and protected health information (like the conditions you have and medications you take). The Hugo platform is not considered a "covered entity" under the Health Insurance Portability and Accountability Act of 1996 (HIPAA); this means that the HIPAA privacy rule does not apply to this platform. The Hugo platform does take all necessary precautions, including industry-standard encryption, to minimize privacy and security risks to your stored personally identifiable information. To learn more about Hugo's commitment to the security and privacy of your data, you can visit the following links: Security Statement (https://hugo.health/security), Privacy Notice (https://hugo.health/privacy-notice), Terms of Service (https://hugo.health/terms-of-service/).

While participating in this study, you will be asked to fill out multiple surveys, which will take some time and maybe inconvenient. Some of the surveys may include questions that you feel uncomfortable answering. If you feel uncomfortable answering any specific survey question, you will be able to skip these questions and continue with the rest of the survey. You may also find it inconvenient to wear the provided wearable device or connect it to your phone.

Although the device that you will receive has been deemed to not be a 'High-Risk Device' by the FDA and is therefore not deemed a medical-grade device, there is the risk of inaccurate measurements. You may also experience skin irritation from the band and device. This risk may be increased by wearing the device improperly; the research associate will show you the best way to wear your device to minimize this risk.

**Benefits**

A possible benefit of this study is, with the use of the Hugo platform, you will have easy access to your medical information collected by Yale-New Haven Health and outside health records if you choose to. Using the provided wearable device, you may also gain additional awareness and information regarding your health and fitness.

**Economic Considerations**

For your time and commitment to this study you will be compensated. As a participant in this study you will be given a wearable device for data collection and will be allowed to keep the device at the end of the study. If all monthly surveys are completed, you will be eligible to receive a minimum of $150 for your time, which includes the value of the device you receive. Payments will be dependent on which device you receive; once your device has been assigned, the research associate will walk you through how, if applicable, you will receive any payments. If you receive a device valued over $150 you will not receive additional compensation but will be able to keep the device at the end of the study. If you receive a device valued under $150, you will receive monthly payments for completing your surveys, ($15 at baseline, $10 for each monthly survey months 1-5, and $15 for the surveys at 6 months), and will also be able to keep the device provided to you.

If applicable, you will receive payments for your participation in this study via a Visa pre-paid card. As surveys are completed, the balance will be automatically credited to your account which is linked to your email address. You will receive payments every 30 days, as long as you are responding to the study surveys; these payments are provided during just the first 6 months of the study. When these payments are ready, you will receive an email with instructions on how to receive your payment. Your Visa card will then be sent to your email as a digital ecard. You may redeem your payments at any time once you have an available balance.

This study will not affect your health care costs in any way. You will still be responsible for any costs associated with any routine follow-ups or doctor visits, and there will be no additional follow-ups or doctor visits scheduled as part of this study. You will still be responsible for any co-pay required by your insurance company for standard treatments. You are also responsible for data charges that may be incurred for utilizing the online features of Hugo or wearable device applications when not connected to Wi-Fi.

**Treatment Alternatives/Alternatives**

If you decide not to participate in this study, you will still have access to medical care and your medical records as you would normally. You may decline to participate in the study for any reason without affecting your medical care.

**Confidentiality and Privacy**

Data transferred as part of this study will be sent using a secure, encrypted, and password-protected system. Identifiable information gathered for this study will be kept confidential and disclosed only with your permission. When the results of the research are published or presented, no information will be included that would reveal your identity, unless your specific consent for this activity is obtained.

A description of this study is available on http://www.ClinicalTrials.gov, as required by U.S. Law. This Web site will not include information that can identify you. At most, the Web site will include a summary of the results. You can search this Web site at any time.

The information about your health that will be collected in this study includes:

- Electronic medical records from all health systems that you import into Hugo, including from Yale-New Haven Health System
- Records collected from any pharmacies that you connect and import into Hugo
- Data collected from the provided wearable and iPhone health application(s) during the 12-month follow-up period
- Mobile questionnaires to which you respond
- Medicare claims data that you import into Hugo using CMS Blue Button
- Records about phone calls or emails made as part of this research

The data collected in your Hugo account, including data from any portals you connect and responses to any questionnaires you complete, will not be transferred back to your medical record. This means that your doctors will not see your responses to the study questionnaires or the information from your wearable device.

Information about you and your health which might identify you may be used by or given to:

1. Representatives from Yale University, the Yale Human Research Protection Program, and the Yale Human Investigation Committee (the committee that reviews, approves, and monitors research on human subjects), who are responsible for ensuring research compliance. These individuals are required to keep all information confidential.
2. The Principal Investigators, at both Yale University and the Mayo Clinic, along with other research staff and collaborators who are assisting with this study

3. Hugo Health, the company that owns the platform, in accordance with its Privacy Policy

All health care providers subject to HIPAA (Health Insurance Portability and Accountability Act) are required to protect the privacy of your information. The research staff at the Yale School of Medicine are required to comply with HIPAA and to ensure the confidentiality of your information. Some of the individuals or agencies listed above may not be subject to HIPAA and, therefore, may not be required to provide the same type of confidentiality protection. They could use or disclose your information in ways not mentioned in this form. However, to better protect your health information, agreements are in place with these individuals and/or companies that require that they keep your information confidential. In addition, even though Hugo Health is not required to comply with HIPAA, they maintain the highest standards of confidentiality and security of your information and will never share your data beyond this study without your expressed explicit permission as described in their privacy notice provided when you sign up for Hugo.

This authorization to use and disclose your de-identified health information collected during your participation in this study will never expire. There is no set time for destroying the information that will be collected for this study. Identifiers will be removed from the identifiable private information and after such removal, the information you contribute to this study could be used for future research and can be distributed to another investigator for future research studies without additional informed consent from you or a legally authorized representative. Outside investigators will not know who you are. Private information such as your name, birth date or medical record number will not be shared with them.

☐ **By checking this box, I acknowledge my contribution to science and that my de-identified data collected as part of this study may be used in future research without further consent from me.**

If you have any questions about the collection and use of information about you or would like to exercise rights that you may have regarding this information, you should ask your research associate.

This authorization to use and disclose your de-identified health information collected during your participation in this study will never expire.

#### Voluntary Participation and Withdrawal

Participation in this study is voluntary and you are free to choose not to take part in this study. Refusing to participate will involve no penalty or loss of benefits to which you are otherwise entitled (such as your health care outside the study, the payment for your health care, and your health care benefits). However, you will not be able to enroll in this research study and will not receive study benefits as a study participant if you do not allow the use of your information as part of this study. If you do decline to participate, we will ask if you would be willing to answer a short, voluntary questionnaire to better understand why you would not like to be a part of this study.

**Withdrawing from the Study**

If you do become a study participant, you are free to quit and withdraw from this study at any time during its course.

To withdraw from the study, you can contact your research associate to let them know that you would no longer like to take part. The email to do this is. At this time the researcher can help walk you through how to stop sharing data from your Hugo account.

When you withdraw from this study, no new health information identifying you will be gathered after that date. Information that has already been collected will be retained and used as noted above.

Withdrawing from the study will involve no penalty or loss of benefits to which you are otherwise entitled. It will not harm your relationship with your own doctors or with Yale-New Haven Health or the care that you receive.

**Questions**

We have used some technical terms in this form. Please feel free to ask about anything you don't understand and to consider this research and the consent form carefully – as long as you feel is necessary – before you make a decision.



APPROVED BY THE YALE UNIVERSITY IRB 5/16/2021

**Authorization and Permission**

I have read (or someone has read to me) this form and have decided to participate in the study described above. Its general purposes, the particulars of my involvement, and possible hazards and inconveniences have been explained to my satisfaction. My signature also indicates that I have received a copy of this consent form.

By signing this form, I give permission to the researchers to use [and give out] information about me for the purposes described in this form. By refusing to give permission, I understand that I will not be able to participate in this research.

Print Name of Participant:______________________________

Signature:____________________________________

Date:_______________________________________

______________________________________________ ____________________
Signature of Principal Investigator Date
*or*

______________________________________________ ____________________
Signature of Person Obtaining Consent Date

☐ **By checking this box and signing above, I, the study personnel obtaining consent, confirm that the participant has met all of the defined inclusion criteria for this study.**

If after you have signed this form you have any questions about your privacy rights, please contact the Yale Privacy Officer at 203-432-5919.

If you have further questions about this project or if you have a research-related problem, you may contact the research associate at. If you would like to talk with someone other than the researchers to discuss problems, concerns, and questions you may have concerning this research, or to discuss your rights as a research participant, you may contact the Yale Human Investigation Committee at 203-785-4688.
